# The 22q11.2 deletion syndrome from a biopsychosocial perspective: an ICF-based approach

**DOI:** 10.1101/2024.01.10.24301135

**Authors:** Ana Paula Corrêa Cabral, Dafne Dain Gandelman Horovitz, Lidiane Nogueira Santos, Amanda Oliveira de Carvalho Carvalho, Cristina Maria Duarte Wing, Luciana Castaneda, Liane Simon, Carla Trevisan M. Ribeiro

**Affiliations:** National Institute of Women’s, Children’s and Adolescents’ Health Fernandes Figueiras (IFF/Fiocruz), Rio de Janeiro, RJ, Brazil; Federal University of Rio de Janeiro (UFRJ), Rio de Janeiro, RJ, Brazil; Federal Institute of Rio de Janeiro (IFRJ), Rio de Janeiro, RJ, Brazil; MSH Medical School Hamburg, Hamburg, Germany

**Keywords:** 22q11.2 deletion syndrome, velo-cardio-facial syndrome, International Classification of Functioning Disability and Health, Child, functioning

## Abstract

**Purpose:** To describe children with 22q11.2 deletion syndrome functioning from a biopsychosocial perspective, focusing on the impact of children’s health condition from domains to the International Classification of Functioning, Disability and Health (ICF).

**Methods:** A descriptive, cross-sectional case series study with 22q11.2del children. A questionnaire with an ICF checklist for 22q11.2del was completed using a structured interview. The Test of Childhood Language ABFW was used to fill in vocabulary, fluency and language pragmatics questions. The Wechsler Abbreviated Scale of Intelligence (WASI) was used to determine Intelligence Quotient (IQ).

**Results:** Seven participants from 7 to 12 years old, presented some level of IQ impairment. Observed that 22q11.2del children experience significant intellectual, cognitive and speech impairment across ICF Body Function domains. Impairment related to nose and pharynx were found in only one patient. The most relevant categories considered limitations in Activity and Participation component were pertained to producing nonverbal messages, communication, handling stress and social interaction. Family, health professionals and acquaintances were perceived as facilitators in the component Environmental Factors.

**Conclusion:** Children with 22q11.2del have their functioning affected by aspects that go beyond medical diagnosis. A checklist base on ICF for functional profile can contribute to incorporate a biopsychosocial approach.

## Introduction

The 22q11.2 deletion syndrome (22q11.2DS; 22q11.2del) is the most common microdeletion syndrome in humans. Studies show a frequency of 1:992 for deletions of 22q11.2 in low risk pregnancies and of 1:2,000 livebirths [1]. Clinical manifestations of 22q11.2del are highly heterogeneous and can range from mild to severe, and typically include a congenital heart defect (CHD), velopharyngeal insufficiency with or without cleft palate, mild to moderate immune impairment, characteristic facial features, learning disabilities and language difficulties [2]. Impaired cognitive development is present in 70% to 80% of patients, though severe learning disability is rare [3]. This is in part identified by the presence of reduced Intelligence Quotient (IQ) scores [4]. In addition, subjects with a 22q11.2DS deletion show a higher risk for the development of psychiatric problems during adolescence and early adulthood[5].

Furthermore, speech and language difficulties comprise one of the most distressing aspects for parents of children with 22q11.2DS [2].Studies have highlighted that the difficulties in the production of spoken language presented in del22q11.2 seem to exceed the anatomofunctional alterations of the phono articulatory organs[6–8], with difficulties being found in more complex tasks such as in the production of oral narrative, in the task of verbal fluency [9], and other communication skills [10]. This can have a significant impact on functionality.

Despite the prevalence of the syndrome, it frequently goes undiagnosed because of mild or variable phenotypic expression. Diagnosis can be confirmed by fluorescence in situ hybridization (FISH), a cytogenetic test which is not available in many cases due to their high cost. Hence, it is important to identify the common symptoms as well as the functional domains that may be affected in 22q11.2del children to diagnose, plan comprehensive evaluations, rehabilitation therapies, and long-term follow-up [11].

In this context, the World Health Organization created the International Classification of Functioning, Disability and Health (ICF), which serves as a companion to the International Classification of Diseases in order to characterize how health-related conditions affect people’s lives. The ICF focuses on how context affects functioning and the level of disability [12]. In this case, “functioning” does not refer to quantification of ability or symptoms, but rather to how those symptoms affect an individuals’ age-appropriate ability to respond to the demands of daily life. A biopsychosocial framework for comprehensively describing functioning and disability that is provided by the ICF may enhance our understanding of the broader impact of 22q11.2del on the functioning of individual children.

Recent research studies have begun to use the ICF to assess children’s functioning status in the context of their health condition [13–16]. Havstam et al. [13] were the first study for children with cleft lip and palate (CLP) to utilize the ICF to provide an integrated and comprehensive understanding of CLP and how a communication disability may emerge. Additionally, studies have applied the framework of the ICF to consider children’s communication [15]. However, to our knowledge, applications of the ICF related to children with 22q11.2del specifically are lacking.

The ICF framework can provide health professionals not only with the opportunity to value the positive aspects of functioning, as well as the disability aspects. Thus, the purpose of this study was to describe children with 22q11.2del functioning from a biopsychosocial perspective, focusing on the impact of children’s health condition from four major domains of functioning: Body Functions, Body Structures, Activities, and Participation, and Environmental Factors. The hypothesis is that these children have their functioning affected by aspects that go beyond medical diagnosis.

## Methods

We performed a descriptive, quantitative, cross-sectional case series study with seven 22q11.2 deletion syndrome children. Patients were recruited from a high-complexity hospital in Rio de Janeiro, Brazil between February 1^St^, 2019 to February 31^st^, 2020, and they were either inpatients or outpatients. This is a public hospital which offers multi-professional advice, support and treatment to children and youth with physical, cognitive and/or psychiatric disabilities. The research protocol followed the normative guidelines of Resolution No. 466/12 of the National Health Council of the Brazilian Ministry of Health. Local ethics committee approval was obtained prior to start (n.2.970.203) and patient data was kept anonymous. The consent form of all participants was obtained in writing and documented on a specific form signed by the local Ethics Committee.

Participants for this study were children and inclusion criteria for the study were a positive genetic test, FISH microarray for the 22Q11.2 chromosomal deletion and age between 6 and 12 years old. The families of all eligible children were contacted for participation directly in the clinic by phone or by mail. The study participants were invited consecutively to the clinic at the times when the investigator was present. Parents’ informed consent was obtained and signed in person by them and the researcher before the assessments. Children between 6 and 12 years old also signed in person, together with the researchers and their parents, a free and informed consent form.

The most common impairments of 22q11.2del children were explored using the ICF checklist developed in a previous study [16]. A questionnaire with this checklist for 22q11.2del was completed using a structured interview with the children and parents by the investigator’s direct observations, and by information retrieved from medical records. All participants and their parents were interviewed by health professionals trained in the principles of the ICF. Before an interview started, each patient’s medical record sheet was checked and relevant information on socio-demographic variables and diagnoses were extracted. The participants were also instructed to bring updated medical reports and exam results to the interview for additional information. The data was collected by using a patient profile questionnaire containing demographic information, medical diagnoses, and information about other health-related issues, which were recorded using a patient profile questionnaire.

Impairment in 23 items from the Body Functions component and nine from the Body Structure component were investigated by using the ICF checklist for 22q11.2del. We also checked limitations in 44 items from the Activities and Participation component and assessed as barriers or facilitators 28 items from the Environmental Factors component [16].

The answers from the study population were considered as absolute frequencies of impairments/limitations for the ICF components Body Functions, Body Structures and Activities and Participation. As for the Environmental Factors, absolute frequencies of categories were named to be facilitator or barrier. The ICF categories considered not applicable by the participants were marked as “not relevant/not applicable”.

The Test of Childhood language ABFW was used only to fill in vocabulary, fluency and language pragmatics questions in the ICF checklist [17]. We performed an adapted version of the Portuguese Wechsler Abbreviated Scale of Intelligence (WASI)[18 to determine the full-scale, verbal and performance IQ of the children. WASI is a brief instrument used to assess people’s intellectual ability from 6 to 89 years old. It consists of four subtests, two of which are part of a verbal scale (Vocabulary and Similarities), and two of which are part of a performance scale (Block Design and Matrix Reasoning). The scale was performed by a trained psychologist at a previous scheduled time, and it took around forty minutes. Age-adjusted scores were derived and transformed to a score using norms available for each measure. These scores are already age-standardized, where higher scores indicate better performance.

Data was collected manually initially, and then entered, organized, and analyzed using Epi Info software version 7.0. Following that, the data was exported and descriptive statistics (frequency, percentage, mean) were used to describe the studied variables.

## Results

Our analyses revealed that five out of seven of them were boys; only one subject presented with a hearing loss diagnosis; and the population age was from 7 to 12 years old with a mean of 10.28 and a std. deviation of 2.05 (Table 1). The IQ was assessed using WASI by measuring the verbal IQ (VIQ), performance IQ (PIQ) and general cognition full-scale (FSIQ). Table 1 shows the mean performance on WASI scores and comparison of means between participants. Cognitive assessment was completed for six of the seven patients enrolled in the study. Due to scheduling in a different day from the ICF questionnaire application, cognitive assessments were not performed in one patient, who did not reply to phone calls, mails, or text messages; after a period of ten months without any contact the participant was excluded from cognitive testing. We identified that all subjects presented with IQ impairment. In the verbal IQ, one child had an average IQ, one had a low average intellectual disability, two had an IQ in the borderline range, and two had an extremely low verbal IQ. As for the performance IQ, two children had a low average, three had a borderline range, and one had an extremely low performance IQ. Finally, in the full-scale IQ, two children had low average IQ, one had an IQ in the borderline range, and three had an extremely low full-scale IQ. In sum, we found that children with 22q11.2del had significant deficits across all cognitive measures: full scale IQ, verbal IQ, and performance IQ scores.

**Table 1.** Demographic characteristics of 22q11.2DS children.

| Population Profile |  | n |
| --- | --- | --- |
| Age | Mean | 10,28* |
|  | Std. deviation | 2,05* |
| Gender | Female | 2 |
|  | Male | 5 |
| 22q2.1del (FISH) | Yes | 7 |
| Hearing loss | Yes | 1 |
|  | No | 6 |
| WASI<br>Verbal IQ (VIQ) | Average | 1 |
|  | Low Average | 1 |
|  | Borderline | 2 |
|  | Extremely Low | 2 |
| WASI<br>Performance IQ (PIQ) | Borderline | 3 |
|  | Low Average | 2 |
|  | Extremely Low | 1 |
| WASI<br>Full Scale IQ (FSIQ) | Borderline | 1 |
|  | Low Average | 2 |
|  | Extremely Low | 3 |

\*age=numbers related to the mean and standard deviation from all subjects (n)= total number of subjects Source: the author, 2022

At the level of Body Functions and Structures, we identified that impairment related to nose and pharynx were found in only one patient, and none of the children presented with external ear structure impairment. We observed that 22q11.2del children experience significant intellectual, cognitive and speech impairment across ICF Body Function domains (Table 2). This is especially identified in the items related to fluency and rhythm of speech and intellectual functions, where we had the highest number for confirmed impairment (n=5). Further, areas related to personality also showed to affect four out of seven children.

**Table 2.** Body Structures and Functions in children with 22q11.2del.

| Categories | Impairment |  | Not evaluated/<br>Not applicable |
| --- | --- | --- | --- |
|  | Yes | No |  |
| **Body Structures** |  |  |  |
| Structure of brain (s110) | - | 4 (57,14%) | 3 (42,86%) |
| Structure of external ear (s240) | - | 7 (100,00%) | - |
| Structure of middle ear (s250) | - | 6 (85,71%) | 1 (14,29%) |
| Structure of inner ear (s260) | - | 6 (85,71%) | 1 (14,29%) |
| Structure of nose (s310) | 1 (14,29%) | 5 (71,43%) | 1 (14,29%) |
| Structure of mouth (s320) | - | 4 (57,14%) | 3 (42,86%) |
| Structure of pharynx (s330) | 1 (14,29%) | 4 (57,14%) | 2 (28,57%) |
| Structure of larynx (s340) | - | 5 (71,43%) | 2 (28,57%) |
| Structure of head and neck region (s710) | - | 7 (100,00%) | - |
| **Body Functions** |  |  |  |
| Consciousness functions (b110) | 1 (14,29%) | 6 (85,71%) | - |
| Orientation functions (b114) | - | 7 (100,00%) | - |
| Intellectual functions (b117) | 5 (71,43%) | 1 (14,29%) | 1 (14,29%) |
| Global psychosocial functions (b122) | - | 7 (100,00%) | - |
| Temperament and personality functions (b126) | 4 (57,14%) | 1 (14,29%) | 2 (28,57%) |
| Energy and drive functions (b130) | - | 7 (100,00%) | - |
| Attention functions (b140) | - | 7 (100,00%) | - |
| Memory functions (b144) | 1 (14,29%) | 6 (85,71%) | - |
| Psychomotor functions (b147) | - | 7 (100,00%) | - |
| Emotional functions (b152) | - | 7 (100,00%) | - |
| Perceptual functions (b156) | 2 (28,57%) | 5 (71,43%) | - |
| Thought functions (b160) | 3 (42,86%) | 4 (57,14%) | - |
| Higher-level cognitive functions (b164) | 2 (28,57%) | 5 (71,43%) | - |
| Mental functions of language (b167) | - | 7 (100,00%) | - |
| Calculation functions (b172) | 1 (14,29%) | 6 (85,71%) | - |
| Experience of self and time functions (b180) | - | 7 (100,00%) | - |
| Hearing functions (b230) | 1 (14,29%) | 6 (85,71%) | - |
| Voice functions (b310) | 1 (14,29%) | 6 (85,71%) | - |
| Articulation functions (b320) | 7 (100,00%) | - | - |
| Fluency and rhythm of speech functions (b330) | 5 (71,43%) | 2 (28,57%) | - |
| Alternative vocalization functions (b340) | 1 (14,29%) | 6 (85,71%) | - |
| Respiration functions (b440) | 1 (14,29%) | 6 (85,71%) | - |
| Muscle tone functions (b735) | - | 7 (100,00%) | - |

**Source:** the author, 2022

We also observed that all participants presented with a major limitation/ restriction related to Activity and Participation in producing nonverbal messages (table 3). Other restrictions for the children were related to communication and social interaction, where the majority of them (four or more children) showed to have participation limitations. Items such as community life, complex interpersonal interactions, abilities to discuss, having a conversation, communicating with written and spoken messages, as well as in writing, reading, thinking were also pointed out as limitations for the study population (table 3).

**Table 3.** Activity and Participation of children with 22q11.2del.

| Categories | Limitation / Restriction |  | Not evaluated/N ot applicable |
| --- | --- | --- | --- |
|  | Yes | No |  |
| Watching (d110) | - | 7 (100,00%) | - |
| Listening (d115) | - | 7 (100,00%) | - |
| Other purposeful sensing (d120) | - | 7 (100,00%) | - |
| Copying (d130) | - | 7 (100,00%) | - |
| Rehearsing (d135) | 1 (14,29%) | 6 (85,71%) | - |
| Learning to read (d140) | 3 (42,86%) | 4 (57,14%) | - |
| Learning to write (d145) | 3 (42,86%) | 4 (57,14%) | - |
| Learning to calculate (d150) | 2 (28,57%) | 5 (71,43%) | - |
| Acquiring skills (d155) | 1 (14,29%) | 6 (85,71%) | - |
| Focusing attention (d160) | 5 (71,43%) | 2 (28,57%) | - |
| Thinking (d163) | 5 (71,43%) | 2 (28,57%) | - |
| Reading (d166) | 5 (71,43%) | 2 (28,57%) | - |
| Writing (d170) | 5 (71,43%) | 2 (28,57%) | - |
| Calculating (d172) | 2 (28,57%) | 4 (57,14%) | 1 (14,29%) |
| Solving problems (d17) | 4 (57,14%) | 2 (28,57%) | 1 (14,29%) |
| Making decisions (d177) | 3 (42,86%) | 4 (57,14%) | - |
| Undertaking a single task (d210) | 3 (42,86%) | 4 (57,14%) | - |
| Undertaking multiple tasks (d220) | 5 (71,43%) | 2 (28,57%) | - |
| Handling stress and other psych (d240) | 4 (57,14%) | 3 (42,86%) | - |
| Communicating with - receiving - spoken messages (d310) | 4 (57,14%) | 3 (42,86%) | - |
| Communicating with - receiving - nonverbal messages (d315) | 1 (14,29%) | 6 (85,71%) | - |
| Communicating with - receiving - formal sign language messages (d320) | 2 (28,57%) | 5 (71,43%) | - |
| Communicating with - receiving - written messages (d325) | 5 (71,43%) | 2 (28,57%) | - |
| Speaking (d330) | 2 (28,57%) | 5 (71,43%) | - |
| Producing nonverbal messages (d335) | 7 (100,00%) | - | - |
| Producing messages in formal sign language (d340) | 4 (57,14%) | 3 (42,86%) | - |
| Writing messages (d345) | 3 (42,86%) | 4 (57,14%) | - |
| Conversation (d350) | 4 (57,14%) | 3 (42,86%) | - |
| Discussion (d355) | 5 (71,43%) | 2 (28,57%) | - |
| Using communication devices and techniques (d360) | 1 (14,29%) | 6 (85,71%) | - |
| Using transportation (d470) | 1 (14,29%) | 5 (71,43%) | 1 (14,29%) |
| Acquisition of goods and services (d620) | 1 (14,29%) | 5 (71,43%) | 1 (14,29%) |
| Basic interpersonal interactions (d710) | 3 (42,86%) | 4 (57,14%) | - |
| Complex interpersonal interactions (d720) | 4 (57,14%) | 3 (42,86%) | - |
| Formal relationships (d740) | 2 (28,57%) | 5 (71,43%) | - |
| Informal social relationships (d750) | 1 (14,29%) | 6 (85,71%) | - |
| Family relationships (d760) | 1 (14,29%) | 6 (85,71%) | - |
| Informal education (d810) | 4 (57,14%) | 3 (42,86%) | - |
| Preschool education (d815) | 1 (14,29%) | 6 (85,71%) | - |
| School education (d820) | 2 (28,57%) | 5 (71,43%) | - |
| Basic economic transactions (d860) | 2 (28,57%) | 5 (71,43%) | - |
| Community life (d910) | 4 (57,14%) | 3 (42,86%) | - |
| Recreation and leisure (d920) | 1 (14,29%) | 6 (85,71%) | - |
| Human rights (d940) | 3 (42,86%) | 4 (57,14%) | - |

**Source:** the author, 202

Furthermore, we investigated which Environmental Factors related to the physical, social and attitudinal environment in which people live and conduct their lives might be facilitators or barriers to children’s functioning. Within the ICF checklist related to children with 22q11.2del, we found that the Environmental Factors that were considered facilitators for all participants were their immediate family (e310), transportation services, systems, and policies (e540), utility services, systems and policies (e530), media services, and systems and policies (e560) (Table 4). Most of the participants (5 out of 7) also considered facilitators their extended family (e315), acquaintances, peers’ colleagues, neighbors, and community members (e325), as well as personal care providers and personal assistants (e340), health professionals (e355), individual attitudes of immediate family members (e410) and communication services, systems and policies (e535) (Table 4).

**Table 4.** Environmental Factors related to children with 22q11.2del.

| Categories | Facilitator |  | Not evaluated/N ot applicable |
| --- | --- | --- | --- |
|  | Yes | No |  |
| Immediate family (e310) | 7 (100,00%) | - | - |
| Extended family (e315) | 5 (71,43%) | 2 (28,57%) | - |
| Friends (e320) | 2 (28,57%) | 5 (71,43%) | - |
| Acquaintances, peers colleagues, neighbours and community members (e325) | 5 (71,43%) | 2 (28,57%) | - |
| Personal care providers and personal assistants (e340) | 5 (71,43%) | 2 (28,57%) | - |
| Domesticated animals (e350) | 3 (42,86%) | 4 (57,14%) | - |
| Health professionals (e355) | 5 (71,43%) | 2 (28,57%) | - |
| Health-related professionals (e360) | 4 (57,14%) | 3 (42,86%) | - |
| Individual attitudes of immediate family members (e410) | 5 (71,43%) | 2 (28,57%) | - |
| Individual attitudes of extended family members (e415) | 3 (42,86%) | 3 (42,86%) | 1 (14,29%) |
| Individual attitudes of friends (e420) | 2 (28,57%) | 4 (57,14%) | 1 (14,29%) |
| Individual attitudes of acquaintances, peers colleagues, neighbours and community members (e425) | 3 (42,86%) | 4 (57,14%) | - |
| Individual attitudes of personal care providers and personal assistants (e440) | - | 6 (85,71%) | 1 (14,29%) |
| Individual attitudes of health professionals (e450) | - | 6 (85,71%) | 1 (14,29%) |
| Individual attitudes of health-related professionals (e455) | - | 6 (85,71%) | 1 (14,29%) |
| Societal attitudes (e460) | 1 (14,29%) | 5 (71,43%) | 1 (14,29%) |
| Utilities services, systems and policies (e530) | 7 (100,00%) | - | - |
| Communication services, systems and policies (e535) | 5 (71,43%) | 2 (28,57%) | - |
| Transportation services, systems and policies (e540) | 7 (100,00%) | - | - |
| Associations and organizational services, systems and policies (e555) | 4 (57,14%) | 3 (42,86%) | - |
| Media services, systems and policies (e560) | 7 (100,00%) | - | - |
| Social security services, systems and policies (e570) | 3 (42,86%) | 4 (57,14%) | - |
| General social support services, systems and policies (e575) | - | 6 (85,71%) | 1 (14,29%) |
| Health services, systems and policies (e580) | 4 (57,14%) | 3 (42,86%) | - |

**Source:** the author, 2022

We also identified Environmental Factors related to the use of products, none of the participants make use of products and technology for personal use in daily living (e115), only one participant reported use of products and technology for culture and recreation (e140), two reported use of products for communication (e125), and five children use products and technology for education (e130) (data not shown) (table 4).

The main findings (over than 45%) related to the functionality of children and adolescents with 22q11.2del are described in fig 1.

**Fig 1.**
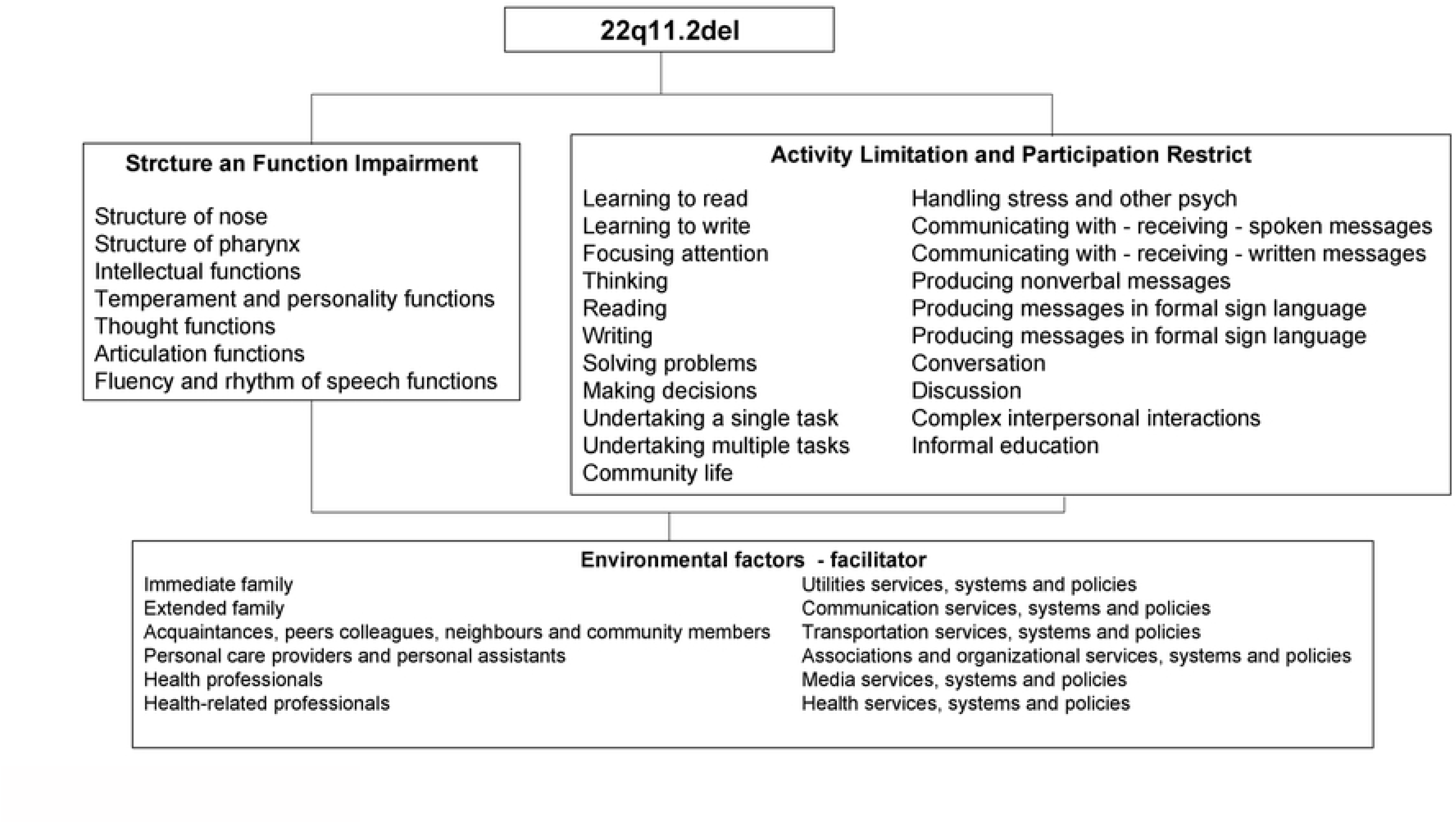
Main findings related to the functionality of children and adolescents with 22q11.2del.

## Discussion

Our findings point out that all four components of the ICF (Body Functions, Body Structures, Activities and Participation, and Environmental Factors) are relevant for children with 22q11.2del, confirming the hypothesis is that these children have their functioning affected by aspects that go beyond medical diagnosis. According to the ICF, impairments are defined as problems in body function or structure. All the variables in this study relating voice and speech to articulation functions, alternative vocalization functions, fluency and rhythm functions are by definition impairments of Body Functions. In agreement, it has been described speech and language disorders in the school aged years such as hypernasality and voice disorders, as well as language impairments related to difficulties with narrative and descriptive language [11,19], which goes along with the findings in our study where fluency and rhythm functions were considered impaired for 5 out of the 7 children interviewed.

Moreover, we found that mental functions were also significantly affected in the children with 22q11.2del, items related to intellectual functions, thought functions, perceptual functions, higher-level cognitive functions, temperament, and personality functions were the most frequently impaired. General mental functions constitute the way the individual reacts in a particular way to situations, including the set of mental characteristics that makes the individual distinct from others [12]. During the interview, 4 children demonstrated variation in emotional reactivity; such behavior was also emphasized by the parents as being frequent in daily routine as they answered the checklist. Our findings are of great importance because impairments in mental functions can signalize risk factors for developing psychiatric disorders. A previous study found similar results when they described 22q11.2del children temperament to be moderately difficult [20]. In line with this, other studies described children with 22q11.2del at increased risk for psychiatric disorders throughout development, especially attention-deficit/hyperactivity disorder (ADHD), anxiety and affective disorders, psychosis, and schizophrenia spectrum disorders [21]. In accordance with our findings, perceptual functions impairment were also found in another study where they reported that 22q11del children were significantly slower to recognize happiness, sadness, and anger (but not fear) on a dynamic emotion task, compared to other children [22]. Such behaviors should be considered since psychiatric disorders are among the most common manifestations of 22q11del and diagnoses characteristically prevalent in childhood include ADHD (30–40%), anxiety (30– 40%) and mood (20–30%) disorders, which increase in prevalence during adolescence [22].

Body Structures impairments were found only in 2 participants, both related to structures involved in voice and speech. This could be related to the specific speech and language disorders that continue into the school aged years [11, 23]. However, the data of the patients in our study are not in line with previous qualitative research when it comes to impairments in Body Structures. In the latter, patients experienced more problems than in this present research. This could be due to a lack of clinical reports on patient’s records and long waiting lists did not make needed and/or new evaluations possible, therefore, in our research, health professionals rated the patients in the Body Structures category according to the medical and professional reports available at the moment, as recommended by the ICF manual [10], therefore complementary exams to fully assess the items from the Body Structures from the ICF checklist for 22q11.2del was not possible and could have impacted the Body Structures impairments assessment. Thus, we cannot confirm the instrument reliability for Body Structures, because the few number of patients having some classifiable impairment in those components prevents a meaningful analysis.

The ICF focuses on how context affects function. “Functioning” does not refer to quantification of symptoms, but rather to how those symptoms affect individuals’ ability to respond to demands of daily life. These domains are related, but independent, the presence of below-average cognitive abilities does not necessarily confer low function, therefore the IQ assessed using the Wechsler Abbreviated Scale of Intelligence (WASI) from which full-scale IQ (FSIQ), verbal IQ (VIQ) and performance IQ (PIQ) was of great worth for understanding 22q11.2del children’s functioning. The level of intelligence in children and adolescents with 22q11.2 deletion syndrome is highly variable [24]. In our study we found that in the FSIQ all children had significant deficit, and 4 out of the 6 children assessed presented with a FSIQ below 70. While divergent cognitive trajectories across age has been reported [25], others suggest some cognitive decline for most 22q11del individuals as they grow into adulthood [26]. This goes in accordance with previous findings where approximately 50% of individuals with 22q112del had intellectual disability (ID), defined by an IQ score of less than 70 [27].

Although the intelligence profile is highly variable in 22q11.2del children, examination of sub-scale scores on the WASI revealed that4 out of the 6 children had lower VIQ scores compared to PIQ scores. Although the magnitude of the difference on average is variable (from 1 to 19 IQ points), it may be worth investigating in future studies if some individuals perhaps speak less or if some aspect of verbal communication is diminished by transmission. These results were also described where a subgroup of children with 22q11.2del during early primary school age showed a discrepancy between verbal abilities and perceptual reasoning abilities, favoring the verbal domain [28].

The most relevant categories considered limitations in Activity and Participation component were pertained to producing nonverbal messages, communication, handling stress and social interaction. Because verbal communication is a major contributor to the foundation of a person’s identity and their social validation, disruptions in one’s speech may result in decreased self-concept. Decrements in self-esteem, in turn, may contribute to feelings of uneasiness during verbal exchanges in social settings [29]. These results indicate a correlation with our findings in Body Structures and Functions, where speech difficulties of children with 22q11.2del were also relevant categories and can potentially be associated with psychological or emotional problems including difficulties with self-esteem, depression, and anxiety [30].

In addition, categories related to the learning process, such as reading, writing, thinking, and focusing attention, were also recorded as a limitation for most of the children. These findings are also described in the literature, where learning disabilities and academic achievement difficulties, as well as deficits in attention and concentration are reported in children with 22q11.2del [9]. In sum, we see that disruption to the Body Functions and Structures of children with 22q11.2del is but one component of functioning as described by the ICF. The interrelated nature of the ICF components suggest that the functioning and disability of an individual emerges from the interaction of one’s Body Functions and Structures with other components of the ICF such as Activity and Participation [12] and this examination of all interactions is necessary for a complete description of functioning in children with 22q11.2del.

Family, health professionals and acquaintances were perceived as facilitators for the 22q11.2del children in the component Environmental Factors. For more than half of the participants health services, systems and policies were also important facilitators. This emphasizes the importance of children’s social network and the influence of a good relationship with health professionals. Although there is a reasonable body of literature describing the IQ, achievement, neurocognitive and behavior abnormalities in children with 22q11del [3,27], there is a lack of published studies that have assessed the role of the environment in functioning in these domains.

Report of use of products were not relevant for this study population, one participant reported to use products and technology for culture and recreation, two reported use of products for communication, and 5 children use products and technology for education, this category contains items like calculator, which was the item cited by all 5 participants. It remains unclear the role of other products and its use since studies that correlate specific environmental factors from the ICF and the 22q11.2del children were not found in the literature.

Studies in general only highlight the deficiencies of structure and function, according to a biomedical approach that is now outdated. The current focus of healthcare, whether clinical or rehabilitation, should be directed towards activity and participation from a biopsychosocial perspective. The use of the ICF can lead to this direction as it considers the subject in its fullest and beyond the health condition itself. Functioning of 22q11.2 deletion syndrome children is an important factor that affects their activity, participation, and quality of life. Although this is a case-series study, thus limiting its generalizability to larger populations of patients, this study can also provide information that allows hypotheses to develop, leading to further advanced studies. An ICF based checklist approach, such as the one used in this research, can be used to identify common profiles of functioning, activities, participation, and environment of 22q11.2 child population. The value of using the ICF in the assessment of children with 22q11.2del is that it enables the health professionals to focus on what should be the goal of intervention: improvement of the child’s functioning.

The findings from our experience on the applicability of an ICF checklist for 22q11.2del shows that ways of operationalizing the ICF can contribute to overcome the biomedical perspective, creating possibilities to incorporate a biopsychosocial approach into the healthcare routine. However, one limitation on our study was that some questions used in the ICF questionnaire for 22q11.2del were not completely clear to the parents, and a previous explanation was needed for better understanding. This should be taken into consideration for improvement in future studies. One limiting factor for this study is also the sample size of 7. In order to draw stronger conclusions, a larger sample size is required. We justify the sample size of the study with the relatively rare nature of 22qdel and the difficulty in recruiting active participants.

Another limitation of the present study is that the clinical records were not enough to evaluate Body Structure and Function, and further evaluation was not always possible. This means that the prevalence of impairments in Body Function and Structure might have been underestimated. Rehabilitation based on the ICF approach focus on a humanized healthcare, where planning interventions are centered in the interaction between an individual’s health conditions, environmental factors and individual factors, designed to remove barriers in several contexts. Therefore, further studies on Body Structure and Function related to Activity and Participation according to the ICF will clarify the impact and impairments of 22q11.2del in children’s functioning.

## Conclusion

Our findings show that children have their functioning affected by aspects that go beyond body Functions and Structures describe in medical diagnosis. Relevant categories related to Activity and Participation limitations were observed, especially in the production of non-verbal messages, communication, stress management and social interaction. Incorporating a biopsychosocial approach is important to expend the assessment of children with 22q11.2del.

This is the first study to use ICF for functional profile of 22q11.2del children and adolescent, therefore, this study represents the first stage in the development of an ICF Core Set in 22q11.2del, hoping that it could be a practical tool to give insight into the wider determinants of the 22q11.2 population and be used in public health and social policy to help address inequalities within the child population. Our experience represents a strong stimulus for extending our sample and verifying the results obtained in this preliminary experience to have a full profile of 22q11.2del functioning based on the ICF. Moreover, our results show that the ICF checklist for children with 22q11.2del fulfils the requirement of a useful, reliable, and valid multidimensional framework for defining domains and describing functioning in children aged 7–12 years.

## Data Availability

All relevant data are within the manuscript and its Supporting Information files.

## Acknowledgments

The authors would like to acknowledge the generous support of the Oswaldo Cruz Foundation, Deutscher Akademischer Austauschdienst (DAAD) and Medical School Hamburg.

## Financial support

IFF/Fiocruz (PIBIC 09412); Deutscher Akademischer Austauschdienst - DAAD (57440919)

## Notes

### Competing Interest Statement

The authors have declared no competing interest.

### Funding Statement

Yes

### Author Declarations

Ethics committee from Instituto Fernandes Figueira/ Fiocruz approval was obtained prior to start (n 3.900.914). The work is registered on Plataforma Brazil (CAEE 26957419.6.0000.5269) Participants signed the Free and Informed Term of Commitment in person.

